# Evaluation of nCoV-QS (MiCo BioMed) for RT-qPCR detection of SARS-CoV-2 from nasopharyngeal samples using CDC FDA EUA qPCR kit as a gold standard: an example of the need of validation studies

**DOI:** 10.1101/2020.05.01.20081034

**Authors:** Byron Freire-Paspuel, Patricio Vega-Mariño, Alberto Velez, Paulina Castillo, Marilyn Cruz, Miguel Angel Garcia-Bereguiain

**Author notes:** corresponding author: Miguel Angel Garcia-Bereguiain.

## Abstract

**Background:** Several qPCR kits are available for SARS-CoV-2 diagnosis, mostly lacking of evaluation due to covid19 emergency.

**Objective:** We evaluated nCoV-QS (MiCo BioMed) kit using CDC kit as gold standard.

**Results:** We found limitations for nCoV-QS: 1) lower sensitivity 2) lack of RNA quality control probe 3) no capacity to quantify viral load.

**Conclussions:** Validation studies should be implemented for any SARS-CoV-2 RT-qPCR commercial kit to prevent unreliable diagnosis.

## Background

Multiple in vitro RT-qPCR diagnosis kits are available on the market for for the detection of SARS-CoV-2. Some of them have received emergency use authorization (EUA) from the U.S. Food & Drug Administration (FDA) while others only report validations made by manufacturers, and in general little is known about their performances using clinical specimens. The CDC designed 2019-nCoV CDC EUA kit (IDT, USA) is based on N1 and N2 probes to detect SARS-CoV-2 that have received positive evaluation on recent reports (1-3), and and RdRP as an RNA extraction quality control. Other kit avalaible in the market is nCoV-QS (MiCo BioMed; South Corea) that include probes “ORF3a” and “N” probes for SARS-CoV-2 detection but no probe for RNA extraction quality control (4).

## Objective

This study compared the performance in terms of positive percent agreement (PPA) of nCoV-QS (MiCo BioMed; South Corea) and 2019-nCoV CDC EUA kit (IDT, USA) primers and probes for SARS-CoV-2 qPCR diagnosis from nasopharyngeal samples.

## Study design

Fiftyfour (54) clinical specimens (nasopharyngeal swabs collected on 0.5mL TE pH 8 buffer) from patients selected as suspicious for SARS-CoV-2 infection were included on this study during the surveillance in Galapagos Islands started on April 8th 2020. Also, six negative controls (TE pH 8 buffer) were included as control for carryover contamination. Both CoV-QS and 2019-nCoV CDC EUA kits were kindly granted by independent donors to be used at SARS-CoV-2 diagnosis laboratory “LabGal” at “Agencia de Regulacion y Control de la Bioseguridad y Cuarentena para Galapagos” at Puerto Ayora in Galapagos Islands (Ecuador), where we considered this validation necessary to guarantee the sensibility of SARS-CoV-2 during the surveillance.

## Results

Twenty-five (25) samples were tested following an adapted version of the CDC protocol (1) using CFX96 BioRad instrument and PureLink Viral RNA/DNA Mini Kit (Invitrogen, USA) as an alternate RNA extraction method, and also interpreting as positive 3 samples where a probe was positive with Ct<40 and the second one with Ct values up to 41.15 (See Table 1 and 2a). We performed this protocol for both nCoV-QS and 2019-nCoV CDC EUA primers and probes kits. Nine samples were negative for both kits; Sixteen samples were positive for 2019-nCoV CDC EUA (range of Ct values: 23.02-41.15 for N1; 24.08-40.12 for N2), but only ten (PPA 62.5%; p<0.001) of those ones were positive for nCoV-QS (range of Ct values: 28.71-39.98 for ORF3a; 24.48-35.44 for N). Results are detailed on Table 1a. The assay was validated to detect 20 viral RNA copies/uL by using 2019-nCoV N positive control (IDT, USA).

**Table 1.** Performance of nCoV-QS compared to 2019-nCoV CDC EUA for RT-qPCR SARS-CoV-2 diagnosis (% values: PPA)

|  | **CDC Probes**<br>**SARS CoV-2 Positive** | **CDC Probes**<br>**SARS CoV-2 Negative** |
| --- | --- | --- |
| **Veri-Q Probes**<br>**SARS CoV-2 Positive** | 22 (66.7%) | 0 |
| **Veri-Q Probes**<br>**SARS CoV-2 Negative** | 11 | 21 |

**Table 2.** Ct values for nCoV-QS and 2019-nCoV CDC EUA RT-qPCR using CDC adapted protocol for 25 samples (a) and MiCoBioMed protocol for 29 samples (b)

| Sample | Conc.<br>[copies/μL] | CT (2019-nCoV CDC EUA) | Result<br>(2019-nCoV CDC<br>EUA) | CT (nCoV-QS) | Result<br>(nCoV-QS) |
| --- | --- | --- | --- | --- | --- |

| Sample | Conc.<br>[copies/μL] | CT (2019-nCoV CDC EUA) |  |  | Result<br>(2019-nCoV CDC<br>EUA) | CT (nCoV-QS) |  | Result<br>(nCoV-QS) |
| --- | --- | --- | --- | --- | --- | --- | --- | --- |
|  |  | N1 | N2 | RP |  | ORF3a | N |  |
| 1 | 193453,3 | 23,02 | 24,8 | 25,04 | Positive | 28,72 | 24,48 | Positive |
| 2 | 151418,7 | 23,34 | 25,16 | 22,69 | Positive | 32,19 | 24,63 | Positive |
| 3 | 28204,0 | 25,53 | 28,02 | 26,11 | Positive | 30,78 | 27,3 | Positive |
| 4 | 7157,3 | 27,31 | 29,82 | 25,43 | Positive | 34,76 | 30,31 | Positive |
| 5 | 699,3 | 30,34 | 33,23 | 25,32 | Positive | 37,29 | 32,25 | Positive |
| 6 | 684,1 | 30,37 | 33,3 | 25,29 | Positive | N/A | 33,58 | Positive |
| 7 | 657,4 | 30,42 | 32,37 | 29,23 | Positive | 38,26 | 32,16 | Positive |
| 8 | 261,6 | 31,62 | 33,76 | 27,32 | Positive | 39,88 | 33,67 | Positive |
| 9 | 236,5 | 31,75 | 34,4 | 21,73 | Positive | 39,98 | 34,24 | Positive |
| 10 | 162,4 | 32,24 | 34,87 | 23,17 | Positive | 30,04 | 35,44 | Positive |
| 11 | 15,2 | 35,33 | 37,47 | 26,37 | Positive | N/A | N/A | Negative |
| 12 | 9,0 | 36,01 | 40,09 | 24,87 | Positive | N/A | N/A | Negative |
| 13 | 2,8 | 37,53 | 39,62 | 25,27 | Positive | N/A | N/A | Negative |
| 14 | 1,7 | 38,2 | 40,12 | 26,97 | Positive | N/A | N/A | Negative |
| 15 | 1,2 | 38,58 | 39,99 | 27,74 | Positive | N/A | N/A | Negative |
| 16 | 0,2 | 41,15 | 39,51 | 27,98 | Positive | N/A | N/A | Negative |
| 17 | N/A | N/A | N/A | 23,11 | Negative | N/A | N/A | Negative |
| 18 | N/A | N/A | N/A | 26,6 | Negative | N/A | N/A | Negative |
| 19 | N/A | N/A | N/A | 25,56 | Negative | N/A | N/A | Negative |
| 20 | N/A | N/A | N/A | 25,12 | Negative | N/A | N/A | Negative |
| 21 | N/A | N/A | N/A | 25,26 | Negative | N/A | N/A | Negative |
| 22 | N/A | N/A | N/A | 27,63 | Negative | N/A | N/A | Negative |
| 23 | N/A | N/A | N/A | 26,35 | Negative | N/A | N/A | Negative |
| 24 | N/A | N/A | N/A | 25,6 | Negative | N/A | N/A | Negative |
| 25 | N/A | N/A | N/A | 27,77 | Negative | N/A | N/A | Negative |

|  |  | N1 | N2 | RP |  | ORF3a | N |  |
| --- | --- | --- | --- | --- | --- | --- | --- | --- |
| 1 | 883226,7 | 23,1 | 22,96 | 24,14 | Positive | 27,11 | 24,03 | Positive |
| 2 | 736560,0 | 23,31 | 23,39 | 27,04 | Positive | 27,51 | 24,39 | Positive |
| 3 | 153530,7 | 25,07 | 24,97 | 28,66 | Positive | 29,16 | 26,22 | Positive |
| 4 | 23205,6 | 27,2 | 27,48 | 27,63 | Positive | 31,22 | 28,19 | Positive |
| 5 | 4244,5 | 29,11 | 29,2 | 27,07 | Positive | 33,65 | 29,76 | Positive |
| 6 | 1886,7 | 30,02 | 30,88 | 29,81 | Positive | 34,23 | 30,84 | Positive |
| 7 | 1706,6 | 30,15 | 30,17 | 32,26 | Positive | 32,02 | 30,46 | Positive |
| 8 | 1573,9 | 30,13 | 30,12 | 31,17 | Positive | 32,26 | 30,53 | Positive |
| 9 | 719,5 | 31,1 | 31,2 | 26,25 | Positive | 35,4 | 31,03 | Positive |
| 10 | 176,0 | 32,69 | 32,77 | 24,04 | Positive | 26,45 | 31,87 | Positive |
| 11 | 143,5 | 32,8 | 32,36 | 31,46 | Positive | 35,09 | 32,81 | Positive |
| 12 | 51,6 | 34,07 | 34,51 | 26,56 | Positive | 39,43 | 39,89 | Positive |
| 13 | 40,3 | 34,35 | 35,39 | 24,38 | Positive | N/A | N/A | Negative |
| 14 | 3,3 | 37,17 | 37,43 | 26,51 | Positive | N/A | N/A | Negative |
| 15 | 1,8 | 37,87 | 38,27 | 30,89 | Positive | N/A | N/A | Negative |
| 16 | 1,6 | 37,63 | 37,62 | 29,84 | Positive | N/A | N/A | Negative |
| 17 | 0,6 | 39,05 | 38,8 | 27,27 | Positive | N/A | N/A | Negative |
| 18 | N/A | N/A | N/A | 28,25 | Negative | N/A | N/A | Negative |
| 19 | N/A | N/A | N/A | 27,07 | Negative | N/A | N/A | Negative |
| 20 | N/A | N/A | N/A | 30,45 | Negative | N/A | N/A | Negative |
| 21 | N/A | N/A | N/A | 30,65 | Negative | N/A | N/A | Negative |
| 22 | N/A | N/A | N/A | 27,92 | Negative | N/A | N/A | Negative |
| 23 | N/A | N/A | N/A | 29,59 | Negative | N/A | N/A | Negative |
| 24 | N/A | N/A | N/A | 29,09 | Negative | N/A | N/A | Negative |
| 25 | N/A | N/A | N/A | 30,16 | Negative | N/A | N/A | Negative |
| 26 | N/A | N/A | N/A | 30,19 | Negative | N/A | N/A | Negative |
| 27 | N/A | N/A | N/A | 30,76 | Negative | N/A | N/A | Negative |
| 28 | N/A | N/A | N/A | 31,06 | Negative | N/A | N/A | Negative |
| 29 | N/A | N/A | N/A | 33,76 | Negative | N/A | N/A | Negative |

Twenty-nine (29) samples were tested following instructions manual from MiCo BioMed for nCoV-QS kit (4), using MiCo BioMed One RT-qPCR kit. PureLink Viral RNA/DNA Mini Kit (Invitrogen, USA) was used for RNA extraction. We performed this protocol for both nCoV-QS and 2019-nCoV CDC EUA primers and probes kits. Twelve samples were negative for both kits; Seventeen samples were positive for 2019-nCoV CDC EUA (range of Ct values: 23.1-39.05 for N1; 22.96-38.8 for N2), but only 12 (PPA 70.5%; p<0.001) of those ones were positive for nCoV-QS (range of Ct values: 26.45-39.43 for ORF3a; 24.03-39.89 for N). Results are detailed on Table 1 and 2b. We used CFX96 BioRad to run qPCR but also results were confirmed using Veri-Q PCR316 instrument from MiCo BioMed (4). The assay sensitivity indicated on manufacturers manual (1.8 copies/uL for OFR3a and 4.24 copies/uL for N) could not be validated because positive control concentration was not provided.

In summary, overall PPA for nCoV-QS was 66.7% (22 out of 33 positives samples for 2019-nCoV CDC EUA; p<0.001), and 70.5% and 62.5% for MiCo BioMed and adapted CDC protocols, respectively. Additionally, considering the viral loads calculated following adapted CDC protocol with 2019-nCoV N positive control (IDT, USA), the limit of detection (viral copies/uL) for nCoV-QS kit is much higher than the one indicated at manufacturer’s manual (4).

## Discussion

Although the main limitation of our study is the sample size (54 specimens), our results support that nCoV-QS kit had a significant lower performance in terms on PPA and sensitivity compared to 2019-nCoV CDC EUA. Also, the lack of any probe for RNA extraction quality control like RdRP and the unreported concentration of positive controls provided for the kit that does not allow viral load calculations, are limitations to be considered when using nCoV-QS kit.

Considering the worldwide high demand of reagents for SARS-CoV RT-qPCR diagnosis, supplies shortage is a fact, actually affecting harder to developing countries like Ecuador. Under this scenario, validation studies are helpful to guarantee the quality of the supplies in the market for every country in the world.

## Ethical considerations

All samples have been submitted for routine patient care and diagnostics. Ethical approval for this study was not required since all activities are according to legal provisions defined by the “Comité de Operaciones Especiales Regional de Galápagos” that is leading the Covid19 suerveillance in Galapagos Islands. Written informed consent has been obtained by each patient. All data used in the current study was anonymized prior to being obtained by the authors.

## Data Availability

All relevant data is included with the manuscript.

## Funding

None.

## Authorship contribution statement

All authors contributed to study conceptualization, experimental procedures and revision and approval of final version of the manuscript.

Byron Freire-Paspuel and Miguel Angel Garcí Bereguiain analyzed the data and wrote the manuscript.

## Declaration of Competing Interest

All authors have no conflict of interest to declare.

## Acknowledgements

We thank the medical personnel from “Ministerio de Salud Pública” at Galapagos Islands and the staff from the “Agencia de Regulación y Control de la Bioseguridad y Cuarentena para Galápagos” for their support. We also thank Dr. Ronald Cedeño from OPS/WHO for his work during Covid 19 surveillance in Galapagos Islands. We specially thank Gabriel Iturralde, Oscar Espinosa and Dr Tannya Lozada from “Dirección General de Investigación de la Universidad de Las Américas” for logistic support to make SARS-CoV-2 diagnosis possible in Galapagos Islands.

## Notes

### Competing Interest Statement

The authors have declared no competing interest.

### Funding Statement

No funding

